# Clinical Outcomes of Left Atrial Appendage Occlusion Versus Switch of Direct Oral Anti-coagulant in Atrial Fibrillation

**DOI:** 10.1101/2023.03.29.23287936

**Authors:** Andrew Kei-Yan Ng, Pauline Yeung Ng, April Ip, Raymond Chi-Yan Fung, Shing-Fung Chui, Chung-Wah Siu, Bryan P Yan

## Abstract

**Background:** Left atrial appendage occlusion (LAAO) has emerged as an alternative to oral anti-coagulation therapy for stroke prevention in atrial fibrillation (AF), but data comparing LAAO with direct oral anti-coagulant (DOAC) is sparse.

**Method:** This cohort study compared LAAO (with or without prior anti-coagulation) with a switch of one DOAC to another DOAC. The primary outcome was a composite of all-cause mortality, ischemic stroke and major bleeding.

**Results:** A total of 2,350 patients (874 in the LAAO group and 1,476 in the DOAC switch group) were generated by 1:2 propensity score matching. After a mean follow up of 1052 ± 694 days, the primary outcome developed in 215 (24.6%) patients in the LAAO group and in 335 (22.7%) patients in the DOAC switch group (hazard ratio [HR], 0.94; 95% confidence interval [CI], 0.80 to 1.12; P=0.516). The LAAO group had a lower all-cause mortality (HR, 0.49; 95% CI, 0.39 to 0.60; P<0.001) and cardiovascular mortality (HR, 0.49; 95% CI, 0.32 to 0.73; P<0.001), but similar risk of ischemic stroke (HR, 0.83; 95% CI, 0.63 to 1.10; P=0.194). The major bleeding risk was similar overall (HR, 1.18; 95% CI, 0.94 to 1.48, P=0.150), but was lower in the LAAO group after 6 months (HR 0.71; 95% CI 0.51 to 0.97; P=0.032).

**Conclusions:** LAAO conferred a similar risk of composite outcome of all-cause mortality, ischemic stroke and major bleeding, as compared with DOAC switch. The risks of all-cause mortality and cardiovascular mortality were lower with LAAO.

**CLINICAL PERSPECTIVE:** *What Is New?:*

- Data comparing left atrial appendage occlusion (LAAO) with direct oral anti-coagulant (DOAC) in patient with atrial fibrillation (AF) was sparse.
- LAAO conferred a similar risk of composite outcome of all-cause mortality, ischemic stroke and major bleeding, as compared with switch of DOAC in patients with AF and intolerant to at least one anti-coagulant.
- The risks of all-cause mortality and cardiovascular mortality were halved with LAAO.

*What Are the Clinical Implications?:*

- This study highlights the potential role of LAAO as a superior alternative to trying another DOAC for patients with AF and intolerant to at least one DOAC.
- Bleeding events after 6 months post LAAO were significantly reduced by 30% in the LAAO group as compared with the DOAC switch group, coinciding with the de-escalation in anti-thrombotic therapy.
- Reduction in bleeding during the first 6 months post LAAO represents a potential opportunity to further improve outcomes after LAAO.

## Background

Atrial fibrillation (AF) is an increasing epidemic and public health challenge, affecting more than 37 million patients (0.51% of world population) and with a 33% increase during the last 20 years.^1^ AF increases the risk of ischemic stroke by five-fold and contributes in one-fourth of all ischemic strokes through thromboembolic mechanism.^2,3^ Given their superior efficacy and/or safety compared with vitamin K antagonist,^4^ direct oral anti-coagulants (DOAC) is now the preferred option for stroke prevention in non-valvular AF.^5,6^

Since more than 90% of left atrial thrombus is formed within the left atrial appendage,^7^ percutaneous left atrial appendage occlusion (LAAO) emerged as an alternative that may obliviate the bleeding risk conferred by anti-coagulation. Due to historical reason, the two landmark trials for LAAO used vitamin K antagonist warfarin, instead of DOAC, in the comparator arm.^8-10^ The only randomized trial comparing LAAO with DOAC was PRAGUE-17 (Left Atrial Appendage Closure vs. Novel Anticoagulation Agents in Atrial Fibrillation), which showed non-inferiority with regards to composite ischemic and bleeding outcomes but with a limited sample size.^11^ Other retrospective studies have compared LAAO with incident DOAC usage.^12,13^ This comparison may not be totally fair because LAAO, recommended as a second line therapy, is generally considered in those with adverse effects from DOAC. Hence the value of LAAO in comparison to DOAC remains unclear. In view of the limited data, we sought to investigate the clinical outcomes of LAAO versus a switch from one DOAC to another DOAC in patients with non-valvular AF.

**Methods**

The data that support the findings of this study are available from the corresponding author upon reasonable request.

### Study Population and Design

This is a cohort study of AF patients undergoing LAAO in all eight public hospitals that performed the procedure in Hong Kong compared with a propensity score–matched control cohort of AF patients with a first switch from one DOAC (dabigatran, rivaroxaban, apixaban, edoxaban) to another DOAC between January 1, 2012 and December 31, 2020. Patients were identified and their baseline characteristics, exposures and outcomes were retrieved from the Clinical Data and Analysis Reporting System (CDARS). CDARS is a territorial-wide electronic health record managed by the Hospital Authority of Hong Kong, which provides hospital care to approximately 90% of the local population as public health service. CDARS records all essential clinical information including patients’ demographics, hospitalisation, visits to out-patient clinics and emergency services, diagnoses, laboratory results, procedures, prescriptions, dispensing of medications and death. Patients were anonymized while clinical data can be retrieved from CDARS with a unique, anonymous patient identifier. Many high-quality, population-based studies and multinational pharmacovigilance studies have been conducted based on the data retrieved from CDARS.^14^ The International Classification of Diseases, Ninth Revision (ICD-9), was used for disease coding and previous studies have verified the accuracy of the coding in CDARS with high positive and negative predictive values of more than 90%.^15,16^ Patients’ management and follow up were per routine usual care at the discretion of managing physicians. The study was approved by the Institutional Review Board of the University of Hong Kong/Hospital Authority.

We included all adult patients (18 years of age or older) with paroxysmal, persistent, or permanent non-valvular AF or atrial flutter, CHA2DS2-VASc ≥2, and underwent either LAAO or DOAC switch. The date of LAAO or first prescription of the new DOAC was used as the index date. Patients were excluded if they have any mechanical heart valves, significant mitral stenosis, an indication for anti-coagulation therapy other than AF, any recent stroke or thromboembolic event within 30 days before index date.

### Definitions of Exposure and Outcome Variables

The primary outcome was a composite of all-cause mortality, ischemic stroke and major bleeding. The secondary outcomes were individual components of the primary outcome, cardiovascular mortality. Major bleeding was defined as a composite of fatal bleeding event, intracranial or intraocular bleeding, bleeding necessitating transfusion, or bleeding that caused a drop in hemoglobin of ≥3g/dL, or bleeding that required surgical intervention for control, corresponding to type 3 or type 5 bleeding according to the Bleeding Academic Research Consortium Definition for Bleeding.^17^

### Statistical Analyses

All analyses were performed with prespecified endpoints and statistical methods. We constructed a propensity score that predicted the likelihood of LAAO versus DOAC switch with variables selected based on components of CHA2DS2-VASc, HAS-BLED scores: congestive heart failure, hypertension, age, diabetes mellitus, ischemic stroke or transient ischemic attack, peripheral vascular disease or history of myocardial infarction, female sex, abnormal liver test, estimated glomerular filtration rate (eGFR), history of major bleeding, baseline anemia (hemoglobulin <13g/dL for men and <12g/dL for women), concomitant use of anti-platelet or non-steroidal anti-inflammatory agent, chronic alcoholism.

The study cohort consisted of two comparison groups – “LAAO group” and “DOAC switch group” – generated by 1:2 propensity-score-matching using a caliper of 0.2 times the standard deviation of the logit of the propensity score. We adopted complete case analysis to handle the very rare occurrence of missing variables. Unadjusted analyses were made using chi-square tests for categorical variables and Student’s t-test or Wilcoxon rank-sum tests for continuous variables. Kaplan-Meier survival curves were constructed for study groups.^18^ Cox proportional hazards regression was performed to evaluate the relationship between exposure and clinical outcomes as a time-to-first-event analysis with administrative censoring at 3 years after the index date, loss to follow up or end of study (July 31, 2021). Since anti-thrombotic therapy is typically de-escalated (e.g. cessation of dual anti-platelet therapy) at 6 months after LAAO, we performed additional landmark analysis for events occurring after 6 months.

### Sensitivity analyses

First, we included all patients before propensity score matching, and used cox regression model to evaluate the primary and secondary outcomes, using the same set of co-variates used in construction of the propensity score model in the primary analysis. Next, since patients with cancer may be less likely to receive LAAO due to perceived limited life expectancy, we repeated the propensity score matching and primary analysis in which all patients with a history of cancer at baseline were excluded, similar to previous study.^13^ Next, since patients undergoing LAAO or DOAC switch due to a prior bleeding event may differ from those without such event, we repeated the primary analysis restricted to patients with a prior history of bleeding.

Data management and statistical analyses were performed in Stata software, version 16 (StataCorp, College Station, Texas). A two-tailed P value of less than 0.05 was considered statistically significant.

## Results

### Patients and Characteristics

Between January 2012 and December 2020, a total of 8,647 patients were considered for inclusion: 1375 (16.0%) were excluded due to any of the following exclusion criteria – thromboembolism within 30 days prior to index date, intracranial hemorrhage within 30 days prior to index date, having an indication for anti-coagulation other than AF, CHA2DS2-VASc score <2. Of the remaining 7,074 patients analysed, 2 were excluded due to missing value (Figure 1). Only 1 (0.04%) patient was lost to follow up, who was in the LAAO group. Baseline characteristics of all patients before propensity score matching is shown in Table S1 of the Supplementary Appendix. A total of 2,350 patients (874 in the LAAO group and 1,476 in the DOAC switch group) were generated by 1:2 propensity score matching. The mean age was 75.9 ± 8.7, in which 1,001 (42.6%) were women. The two groups were well balanced in the bassline characteristics with standardized differences less than 0.1 for all variables except for sex, prior PCI, prior bleeding event and drugs, as shown in Table 1. The most common indication for LAAO was a high HAS-BLED score ≥3 (93.2%) including more than 40% with a history of prior bleeding event (Table S2 in the Supplementary Appendix). In the LAAO group, 642 (73%) received follow up imaging study of left atrial appendage, most commonly with trans-esophageal echo.

**Figure 1.**
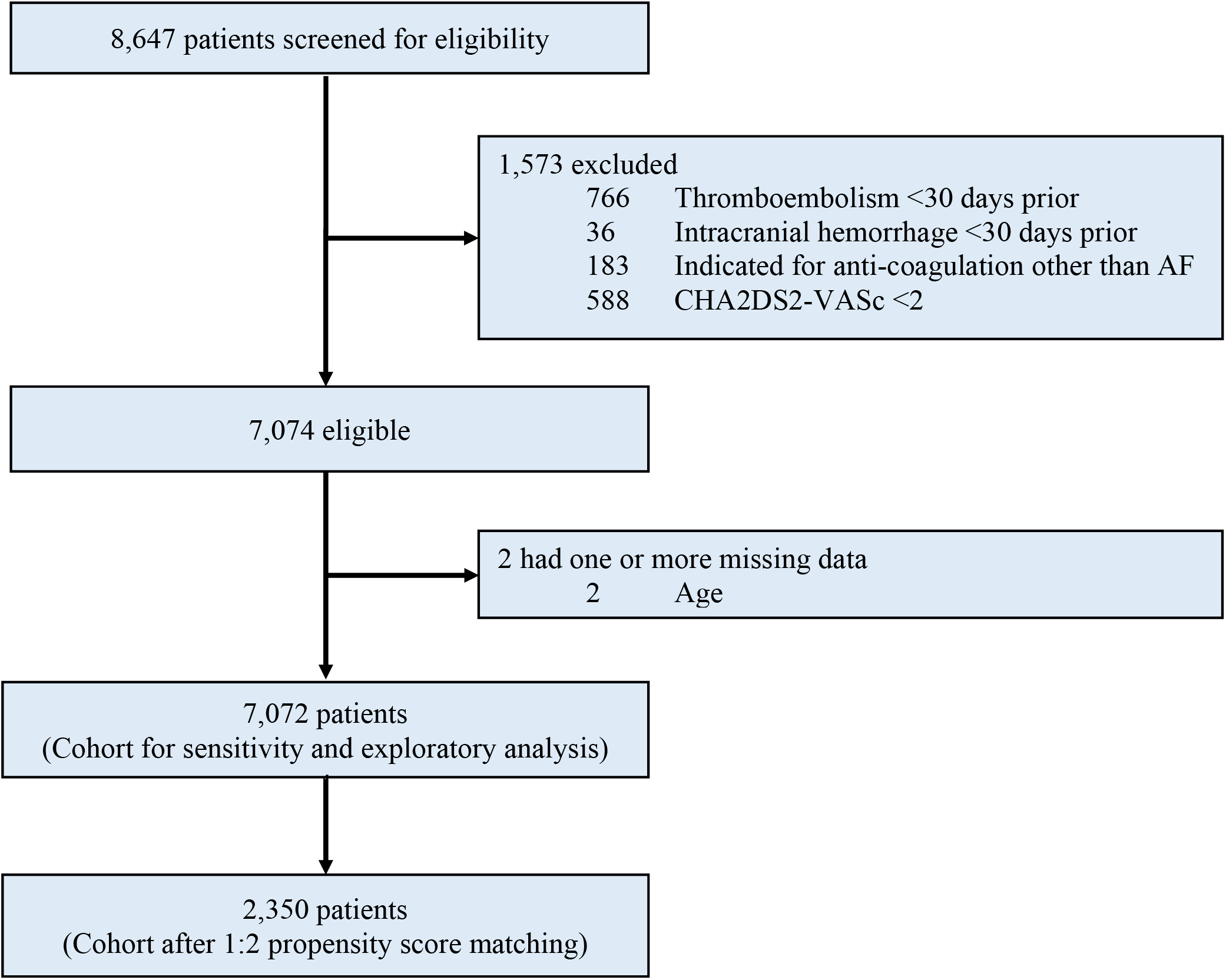
Study profile.* eGFR, estimated glomerular filtration rate; PCI, percutaneous coronary intervention.

**Table 1.**
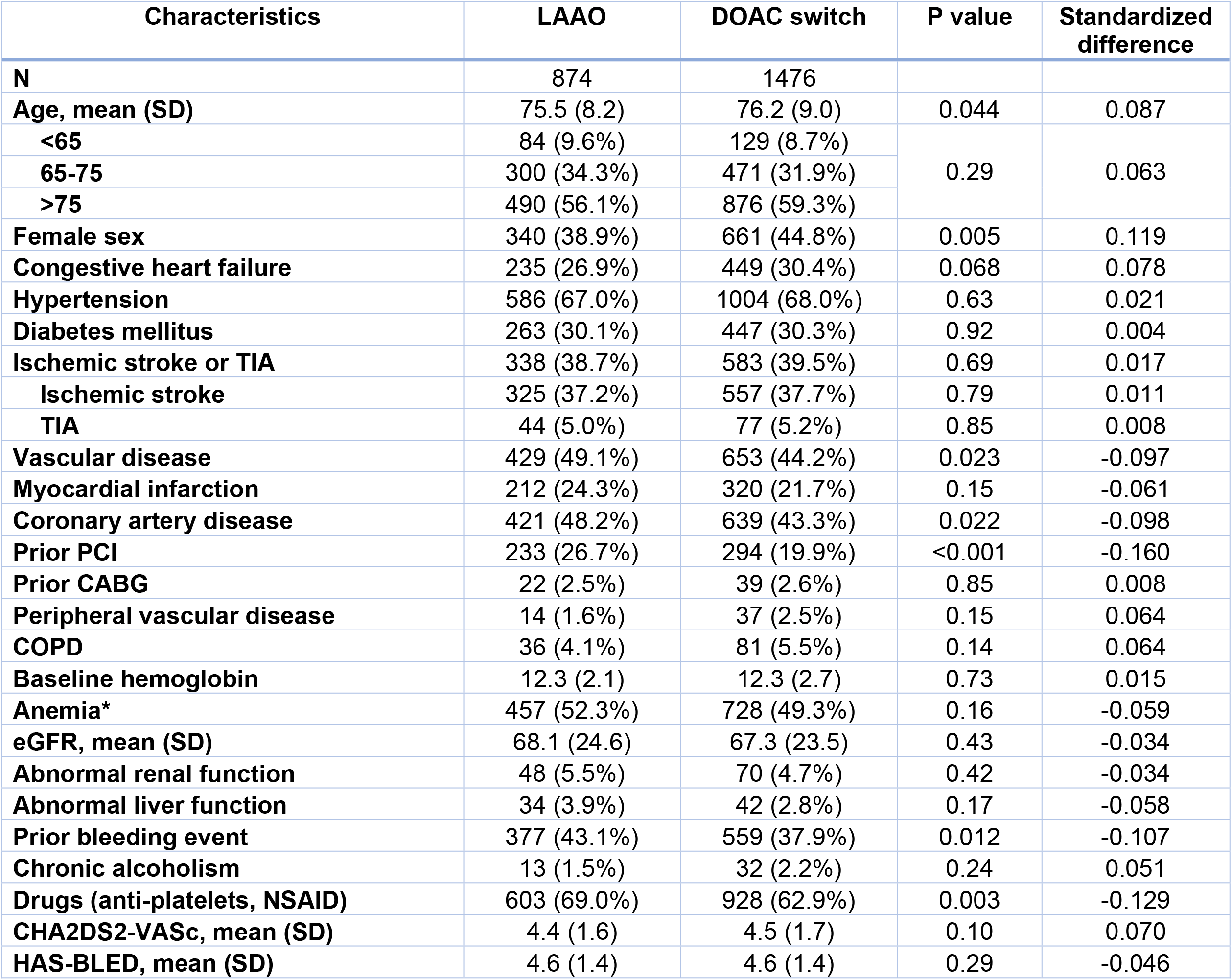
Baseline characteristics of patients. Abbreviations: LAAO, left atrial appendage occlusion; DOAC, direct oral anti-coagulant; SD, standard deviation; TIA, transient ischemic attack; PCI, percutaneous coronary intervention; CABG, coronary artery bypass surgery; COPD, chronic obstructive pulmonary disease; eGFR, estimated glomerular filtration rate *Anemia: Hemoglobin <13g/dL for men, <12g/dL for women.

| Characteristics | LAAO | DOAC switch | P value | Standardized difference |
| --- | --- | --- | --- | --- |
| **N** | 874 | 1476 |  |  |
| **Age, mean (SD)** | 75.5 (8.2) | 76.2 (9.0) | 0.044 | 0.087 |
| **<65** | 84 (9.6%) | 129 (8.7%) | 0.29 | 0.063 |
| **65-75** | 300 (34.3%) | 471 (31.9%) |  |  |
| **>75** | 490 (56.1%) | 876 (59.3%) |  |  |
| **Female sex** | 340 (38.9%) | 661 (44.8%) | 0.005 | 0.119 |
| **Congestive heart failure** | 235 (26.9%) | 449 (30.4%) | 0.068 | 0.078 |
| **Hypertension** | 586 (67.0%) | 1004 (68.0%) | 0.63 | 0.021 |
| **Diabetes mellitus** | 263 (30.1%) | 447 (30.3%) | 0.92 | 0.004 |
| **Ischemic stroke or TIA** | 338 (38.7%) | 583 (39.5%) | 0.69 | 0.017 |
| **Ischemic stroke** | 325 (37.2%) | 557 (37.7%) | 0.79 | 0.011 |
| **TIA** | 44 (5.0%) | 77 (5.2%) | 0.85 | 0.008 |
| **Vascular disease** | 429 (49.1%) | 653 (44.2%) | 0.023 | -0.097 |
| **Myocardial infarction** | 212 (24.3%) | 320 (21.7%) | 0.15 | -0.061 |
| **Coronary artery disease** | 421 (48.2%) | 639 (43.3%) | 0.022 | -0.098 |
| **Prior PCI** | 233 (26.7%) | 294 (19.9%) | <0.001 | -0.160 |
| **Prior CABG** | 22 (2.5%) | 39 (2.6%) | 0.85 | 0.008 |
| **Peripheral vascular disease** | 14 (1.6%) | 37 (2.5%) | 0.15 | 0.064 |
| **COPD** | 36 (4.1%) | 81 (5.5%) | 0.14 | 0.064 |
| **Baseline hemoglobin** | 12.3 (2.1) | 12.3 (2.7) | 0.73 | 0.015 |
| **Anemia\*** | 457 (52.3%) | 728 (49.3%) | 0.16 | -0.059 |
| **eGFR, mean (SD)** | 68.1 (24.6) | 67.3 (23.5) | 0.43 | -0.034 |
| **Abnormal renal function** | 48 (5.5%) | 70 (4.7%) | 0.42 | -0.034 |
| **Abnormal liver function** | 34 (3.9%) | 42 (2.8%) | 0.17 | -0.058 |
| **Prior bleeding event** | 377 (43.1%) | 559 (37.9%) | 0.012 | -0.107 |
| **Chronic alcoholism** | 13 (1.5%) | 32 (2.2%) | 0.24 | 0.051 |
| **Drugs (anti-platelets, NSAID)** | 603 (69.0%) | 928 (62.9%) | 0.003 | -0.129 |
| **CHA2DS2-VASc, mean (SD)** | 4.4 (1.6) | 4.5 (1.7) | 0.10 | 0.070 |
| **HAS-BLED, mean (SD)** | 4.6 (1.4) | 4.6 (1.4) | 0.29 | -0.046 |

### Primary outcomes

The mean follow up period was 1052 ± 694 days. In the propensity match cohort, the primary outcome developed in 215 (24.6%) patients in the LAAO group and in 335 (22.6%) patients in the DOAC switch group, corresponding to an annualized risk of 11.5% and 12.8% respectively (hazard ratio [HR], 0.94; 95% confidence interval [CI], 0.80 to 1.12; P=0.516) (Table 2). Figure 2 shows the Kaplan-Meier event curves, which the LAAO group had a steeper curve in the early period but the DOAC group caught up in the later period. Kaplan-Meier event curves with landmark at 6 months were shown in Figure S1 in the Supplementary Appendix.

**Table 2.** Annualized risks and hazard ratios of primary and secondary outcomes.

| **Outcomes** | **LAAO**<br>**(95% CI)** | **DOAC switch**<br>**(95% CI)** | **Hazard ratio**<br>**(95% CI)** | **P value** |
| --- | --- | --- | --- | --- |
|  | Annualized Rate | Annualized Rate |  |  |
| **<u>Primary outcome</u>** |  |  |  |  |
| **All-cause mortality, ischemic stroke or major bleeding** |  |  |  |  |
| **From index date** | 11.5%<br>(10.0 – 13.1%) | 12.8%<br>(11.5 – 14.3%) | 0.94<br>(0.80 – 1.12) | 0.516 |
| **From 6 months** | 7.8%<br>(6.5 – 9.3%) | 10.5%<br>(9.1 – 12.0%) | 0.75<br>(0.60 – 0.95) | 0.009 |
| **<u>Secondary efficacy</u>** |  |  |  |  |
| **All-cause mortality** |  |  |  |  |
| **From index date** | 5.2%<br>(4.3 – 6.3%) | 11.2%<br>(10.1 – 12.5%) | 0.49<br>(0.39 – 0.60) | <0.001 |
| **From 6 months** | 5.1%<br>(4.2 – 6.3%) | 8.6%<br>(7.5 – 9.9%) | 0.60<br>(0.47 – 0.77) | <0.001 |
| **Cardiovascular mortality** |  |  |  |  |
| **From index date** | 1.5%<br>(1.0 – 2.1%) | 3.2%<br>(2.6 – 3.9%) | 0.49<br>(0.32 – 0.73) | <0.001 |
| **From 6 months** | 1.4%<br>(0.9 – 2.0%) | 2.5%<br>(2.0 – 3.3%) | 0.55<br>(0.34 – 0.90) | 0.016 |
| **Ischemic stroke** |  |  |  |  |
| **From index date** | 4.0%<br>(3.2 – 4.9%) | 4.9%<br>(4.2 – 5.8%) | 0.83<br>(0.63 – 1.10) | 0.194 |
| **From 6 months** | 3.5%<br>(2.8 – 4.5%) | 4.0%<br>(3.2 – 5.0%) | 0.89<br>(0.63 – 1.24) | 0.492 |
| **Major bleeding** |  |  |  |  |
| **From index date** | 7.0%<br>(5.9 – 8.3%) | 6.4%<br>(5.5 – 7.4%) | 1.18<br>(0.94 – 1.48) | 0.150 |
| **From 6 months** | 3.8%<br>(2.9 – 4.9%) | 5.4%<br>(4.5 – 6.5%) | 0.71<br>(0.51 – 0.97) | 0.032 |

Abbreviations: LAAO, left atrial appendage occlusion; DOAC, direct oral anti-coagulant;
CI, confidence interval

**Figure 2.**
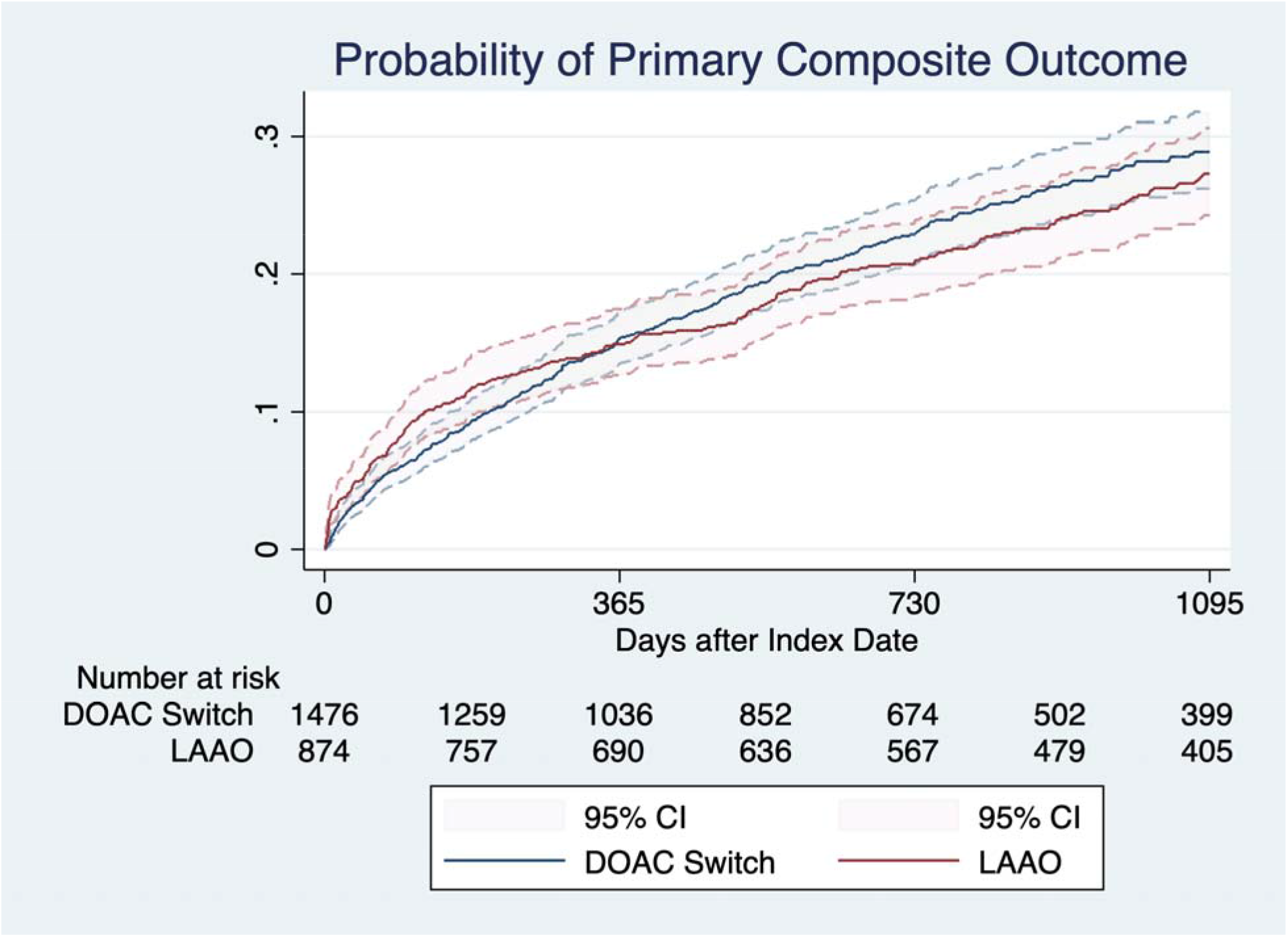
Estimated probabilities of primary composite outcome. The risk of primary composite outcome (all-cause mortality, ischemic stroke, and major bleeding) were similar between groups. Abbreviations: DOAC, direct oral anti-coagulant; LAAO, left atrial appendage occlusion

### Secondary outcomes

The LAAO group had a lower all-cause mortality (HR, 0.49; 95% CI, 0.39 to 0.60; P<0.001) and cardiovascular mortality (HR, 0.49; 95% CI, 0.32 to 0.73; P<0.001) (Table 2). Both groups had similar risk of ischemic stroke (HR, 0.83; 95% CI, 0.63 to 1.10; P=0.194). Overall major bleeding was similar between groups (HR, 1.18; 95% CI, 0.94 to 1.48, P=0.150), but the LAAO group had a lower risk of major bleeding after 6 months (HR 0.71; 95% CI 0.51 to 0.97; P=0.032). Figure 3 shows the Kaplan-Meier event curves for secondary outcomes. Kaplan-Meier event curves with landmark at 6 months were shown in Figure S2-S5 in the Supplementary Appendix.

**Figure 3.**
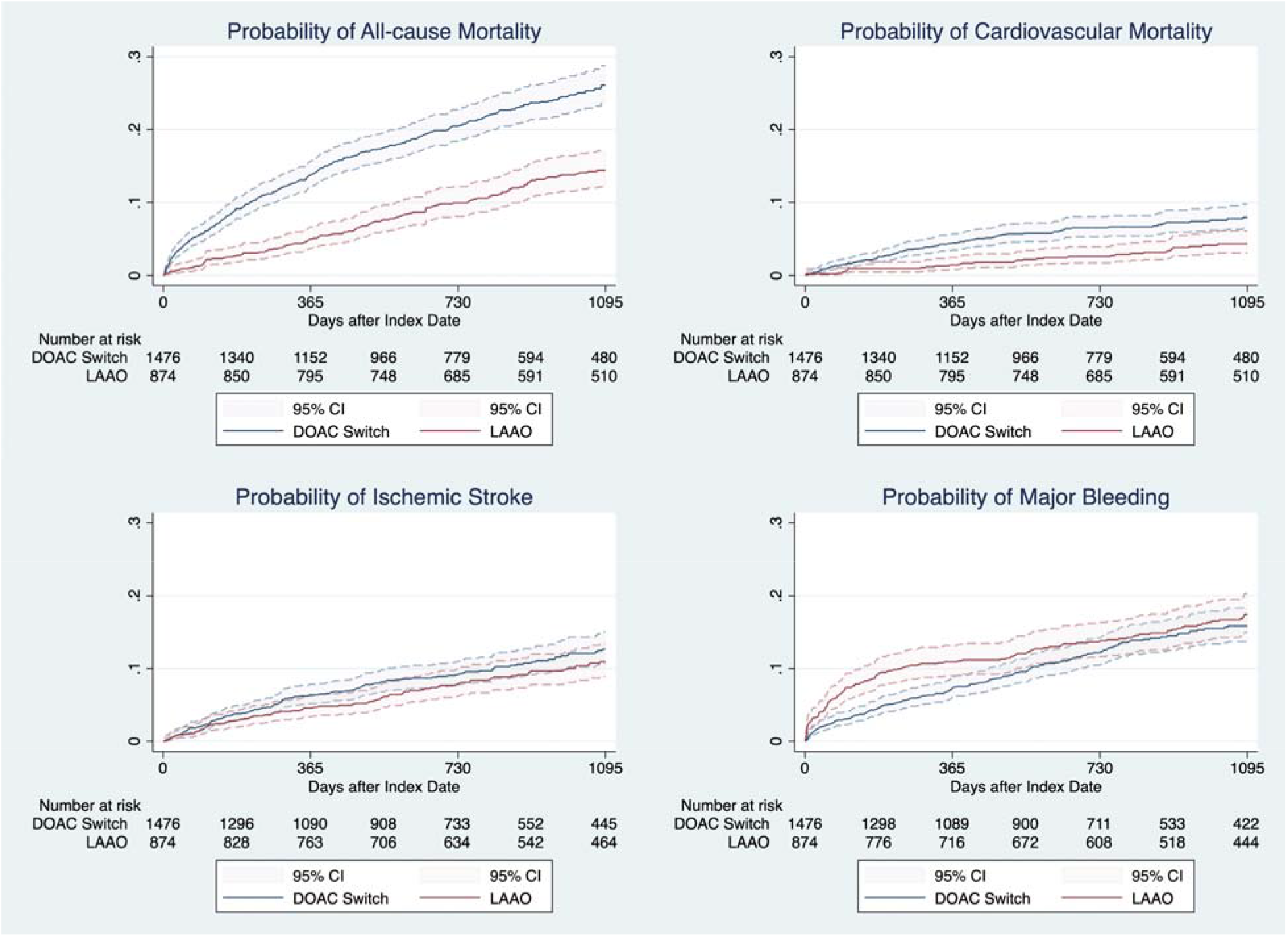
Estimated probabilities of secondary outcomes. Left atrial appendage occlusion group had lower risks of all-cause mortality and cardiovascular mortality, but similar risks of ischemic stroke and major bleeding.

### Sensitivity analyses

Using Cox regression model to assess the outcomes in all patients before propensity score matching, the primary outcome occurred in similar frequency in both groups (HR, 0.92; 95% CI, 0.78 to 1.08; P=0.31). The LAAO had lower risks of all-cause mortality and cardiovascular mortality, but similar risks of ischemic stroke and major bleeding (Table S3 in the Supplemental Appendix). These results were consistent with the primary analysis. Next, after exclusion of patients with a history of cancer, 784 patients in the LAAO group was propensity score matched with 1,315 patients in the DOAC switch group. The primary outcome occurred in similar frequency in both groups (HR, 0.96; 95% CI, 0.80 to 1.15; P=0.63), and all secondary outcomes were consistent with the primary analysis (Table S3 and S4 in the Supplemental Appendix). Next, after restricting the analysis to patients with a prior history of bleeding, the primary outcome occurred in similar frequency in both groups (HR, 0.95; 95% CI, 0.75 to 1.21; P=0.69) (Table S5 in the Supplemental Appendix). Secondary outcomes were also consistent with the primary analysis.

### Safety outcomes of LAAO and patterns of anti-thrombotic therapy

In the 874 patients underwent LAAO, 22 (2.5%) patients had procedurally related major complications, including 19 (2.2%) with pericardial effusion and/or cardiac tamponade, 7 (0.8%) with vascular complications requiring open or endovascular repair and 1 (0.1%) with device embolization. Approximately half of the patients in the LAAO group were prescribed on anti-coagulation therapy upon hospital discharge (Table 3). The median duration of anti-coagulation therapy in the LAAO group was 74 days (Interquartile range, 42 – 155 days). Almost all (93.9%) were free from anti-coagulation therapy at 6 months, but more than half (58.9%) still remained on dual anti-platelet therapy (Table 3). In the DOAC switch group, a significant portion (17.7%) discontinued anti-coagulation therapy at 6 months.

**Table 3.** Anti-thrombotic therapy at different time points in the study groups.

| **LAAO (n = 874)** | **Upon hospital discharge** | **At 6 months** |
| --- | --- | --- |
| **Anti-coagulation therapy** | 412 (47.1%) | 54 (6.1%) |
| **DOAC** | 220 (25.2%) | 45 (5.2%) |
| **Warfarin** | 192 (22.0%) | 9 (1.0%) |
| **Dual anti-platelet therapy** | 793 (90.7%) | 515 (58.9%) |
| **Aspirin** | 813 (93.0%) | 777 (88.9%) |
| **Clopidogrel** | 844 (96.6%) | 554 (63.4%) |
| **DOAC switch (n = 6198)** | **At index date** | **At 6 months** |
| **Anti-coagulation therapy** | 6198 (100%) | 5098 (82.3%) |
| **DOAC** | 6198 (100%) | 5023 (81.4%) |
| **Warfarin** | 0 (0%) | 9 (1.0%) |
| **Dual anti-platelet therapy** | 566 (9.1%) | 201 (3.2%) |
| **Aspirin** | 982 (15.8%) | 560 (9.0%) |
| **Clopidogrel** | 1978 (31.9%) | 941 (15.2%) |

Abbreviations: LAAO, left atrial appendage occlusion; DOAC, direct oral anti-coagulant

## Discussion

In this cohort of AF patients, LAAO had a similar risk of composite outcome of ischemic stroke, major bleeding or all-cause mortality compared with those treated by DOAC switch. LAAO was associated with lower risks of all-cause mortality and major bleeding after 6 months, whereas the risk of ischemic stroke did not differ between groups.

Currently available data directly comparing LAAO with DOAC are very limited. The PRAQUE-17 study was the only published randomized trial comparing LAAO with DOAC.^11^ It showed non-inferiority of LAAO regards to both ischemic and hemorrhagic events, but it had a relatively low number of patients (n = 402) and total events. Two other observational studies compared LAAO with DOAC using propensity-score matching produced somewhat inconsistent results.^12,13^ Importantly, both studies used DOAC-naïve patients as the comparator, which may have unmeasured difference from patients undergoing LAAO because the current practice is still to reserve LAAO for those with a relative or absolute contra-indication to DOAC. Moreover, these studies compared prevalent AF in the LAAO group versus incident AF in the DOAC group which could confer significant bias. A network meta-analysis of randomized clinical trials indirectly suggested that LAAO was less effective than DOAC in stroke prevention but was associated with a lower rate of bleeding.^19^ However if one restricts the meta-analysis to observation studies, LAAO was consistently associated with reduction in both thromboembolic and hemorrhagic events.^19^ In the current study, we compared LAAO with DOAC switch, which had the advantage of a more levalled comparison since the prevailing role of LAAO is still for patients less suitable for DOAC.^5,6^ Our cohort suggested that LAAO had at least a similar overall efficacy compared with DOAC switch in patients with prevalent AF and intolerance to at least one DOAC, with a potential to reduce mortality and late bleeding.

Although the composite outcome was similar between groups, all-cause and cardiovascular mortality rates were significant lower in the LAAO group. This survival benefit was similarly seen in other observational studies comparing LAAO with DOAC.^12,13^ In addition, in the two landmark randomized studies comparing LAAO with warfarin, similar survival benefit was also observed upon long-term follow up, along with reduction in non-procedural bleeding.^8^ The exact reason that LAAO is associated with lower mortality is likely multifactorial, but bleeding reduction from obliviating the need of anti-coagulation therapy plays an important role. Major bleeding in patients with AF is associated with an 8-fold increase in mortality.^20^ Elevated bleeding risk, as determined by the HAS-BLED score, is also associated with excess mortality.^21^ LAAO was shown to reduce bleeding and long-term mortality in randomized trials.^8,10^ Taken altogether, bleeding reduction appears to be an invaluable opportunity to improve clinical outcomes including survival in patients with AF. Our cohort demonstrated similar findings of reduction in mortality and late (presumably non-procedural) bleeding as observed in randomized trials. Therefore data in our cohort suggested that LAAO may improve survival in those intolerant to at least one DOAC. Since mortality is arguably the most important endpoint in patients’ perspective,^22^ LAAO appears to be an attractive alternative to DOAC in selected patients.

In our cohort, the rate of major bleeding was significantly lower in the LAAO group after 6 months. That is likely due to inclusion of procedure related or anti-thrombotic therapy (e.g. short term oral anti-coagulation or dual anti-platelet therapy) related bleeding in the LAAO group. It has been reported that there is a 6.6% to 14.4% annualized risk of major bleeding for the initial phase after LAAO.^23^ On the other hand, patients on DOAC will need to continue DOAC indefinitely and therefore be exposed to the associated bleeding risk. In line with this, we observed more frequent bleeding events in DOAC switch group during the period after 6 months. These patterns are similarly observed in other randomized trials and retrospective studies.^8,13^ In addition, we observed that a considerable portion (17.7%) of patients in the DOAC switch group discontinued DOAC at 6 months, similar to the reported figures in randomized trials for DOAC (21% to 33% on long-term follow up).^24,25^ As a one-off procedure, LAAO may offer superior protection against thromboembolism in this regard. We expect that the benefits of LAAO may accrue over time, but more long-term data and dedicated time-varying outcome analysis is required to prove this conjecture. Studies have also been performed to identify the optimal anti-thrombotic regimen after LAAO, but a conclusion is yet to be established.^23,26^

There has been an emerging trend to adopt a more minimalistic anti-thrombotic strategy after LAAO with dual or even single anti-platelet therapy. In a prospective registry of 1025 patients in Europe and other countries, two-thirds received anti-platelet therapy without anti-coagulation therapy after LAAO.^27^ The registry data suggested that early cessation or omission of anti-coagulation therapy was associated with lower bleeding risk without excess in thromboembolism. In our cohort, nearly half of the patients undergoing LAAO received anti-coagulation upon discharge and the bleeding risk was highest in the first 6 months. Hence this would represent an opportunity to reduce bleeding and hence overall outcomes in patients undergoing LAAO.

This study has some limitations. First, the observational nature of the study conferred risks of unmeasured confounding and bias. Using propensity score matching, we had adjusted extensively for potential confounders that may affect choice of LAAO or DOAC switch, and the findings were consistent in multiple sensitivity analyses. Nonetheless, patients being referred for intervention may have at least a reasonable health status and expected longevity. Second, we only included patients who had some degree of contraindication to anti-coagulation therapy. Our findings may not directly apply to patients who are good candidates to DOAC. Third, information on function outcomes of stroke or major bleeding were unknown to us. Fourth, the LAAO group had more early bleeding events while the DOAC switch group had more late bleeding events. It is uncertain that whether there is any clinical or prognostic difference in the different timing of bleeding. In addition, the landmark analysis at 6 months after index date were not adjusted for multiple comparison and should only be considered exploratory.

## Conclusion

In patients with non-valvular AF, LAAO conferred a similar risk of composite outcome of all-cause mortality, ischemic stroke and major bleeding, as compared with DOAC switch. The risks of all-cause mortality and cardiovascular mortality were lower with LAAO.

## Data Availability

The data could be uploaded for review if requested.

## Non-standard Abbreviations and Acronyms

AF: Atrial fibrillation
CDARS: Clinical Data and Analysis Reporting System
CI: Confidence interval
DOAC: Direct oral anti-coagulant(s)
eGFR: Estimated glomerular filtration rate
HR: Hazard ratio
LAAO: Left atrial appendage occlusion

## Contributors

AKN, CWS and BPY was responsible for the conception and design of the study. AKN analyzed the data collected by AI. AKN interpreted the data. AKN and PYN drafted the manuscript. All authors revised and approved the final manuscript, and are accountable for the accuracy and integrity of the work.

